# Pharmaceutical and Non-Pharmaceutical Interventions for Controlling the COVID-19 Pandemic

**DOI:** 10.1101/2023.03.31.23288023

**Authors:** Jeta Molla, Suzan Farhang-Sardroodi, Iain R Moyles, Jane M Heffernan

## Abstract

Disease spread can be affected by pharmaceutical (such as vaccination) and non-pharmaceutical interventions (such as physical distancing, mask-wearing, and contact tracing). Understanding the relationship between disease dynamics and human behavior is a significant factor to controlling infections. In this work, we propose a compartmental epidemiological model for studying how the infection dynamics of COVID-19 evolves for people with different levels of social distancing, natural immunity, and vaccine-induced immunity. Our model recreates the transmission dynamics of COVID-19 in Ontario up to December 2021. Our results indicate that people change their behaviour based on the disease dynamics and mitigation measures. Specifically, they adapt more protective behaviour when the number of infections is high and social distancing measures are in effect, and they recommence their activities when vaccination coverage is high and relaxation measures are introduced. We demonstrate that waning of infection and vaccine-induced immunity are important for reproducing disease transmission in Fall 2021.

## 1 Introduction

Coronavirus disease 2019 (COVID-19) has been a global challenge leading to millions of infections and thousands of deaths globally. Before the availability of vaccines, most countries relied solely on the implementation of a range of non-pharmaceutical interventions (NPIs) such as partial closings of business, lock-downs, and mask-wearing to curb the spread of SARS-CoV-2 and avoid overburdening healthcare systems [1–3]. With the development of COVID-19 vaccines, policy makers started vaccination campaigns with the aim to protect individuals and relax NPIs. Vaccines became the most important intervention for mitigating disease severity and spread, allowing the return of social and economic activities [4–8].

Human behaviour plays an important role on the efforts to control the transmission of the COVID-19 virus, since the effectiveness of mitigation measures depends on NPI compliance and vaccine acceptance. People are most likely to adapt protective behaviour when mortality or the perception of risk is high, and resume normal life as the perceived risk declines [9–11]. Hence, it is crucial to consider the effects of behaviour change over time so that the design of effective infection mitigation policies can be achieved. Since the onset of the COVID-19 pandemic, many studies have developed mathematical models to describe the dynamics of transmission of the disease [12–14]. Many of the proposed models are extensions of the classical Kermack-McKendrick Susceptible-Infectious-Recovered (SIR) epidemic model [15], which predicts the number of individuals who are susceptible to infection, actively infected, or have recovered from infections at any given time [16]. Several studies have extended the SIR model by considering additional compartments to account for asymptomatic cases, hospitalizations, quarantine, vaccination, disease induced death and /or heterogeneity of the population [17]. These epidemic models can also be coupled with models describing behaviors that are affected by and affect the disease transmission dynamics [18, 19]. Some the proposed COVID-19 compartmental models have considered how individuals respond to the disease dynamics and how the disease dynamics are affected by these behavioural responses [20–30].

In this study, we extend a compartmental SEPIR model first published by Moyles et al. [20]. The model divides the population into five possible disease states: Susceptible (*S*), Exposed (*E*), Presymptomatic (*P*), Infected (*I*) (both symptomatic *I*_*S*_ and asymptomatic *I*_*A*_), and Recovered (*R*). It also includes three classes of social distancing over each disease state. Additionally, infections are delineated into those that are known and unknown. The model was used to study the first several months of the COVID-19 pandemic, and NPI compliance in Ontario, Canada. However, Moyles et al. [20] did not include vaccination, waning immunity, or viral variants as these were concerns after publication of their work. The main purpose of our study is to adapt their model to include vaccination, which confers some immunity to the disease, and waning from all sources of immunity. The effects of waning immunity have been incorporated in some epidemiological models of the COVID-19 pandemic [8,31–35]. Furthermore, we extend the model to include variants of concern by allowing modification of the transmissibility of the disease over time. We do not include the Omicron variant in our study since data acquisition became more difficult as governments reduced testing and started lifting NPIs.

Since vaccination is imperfect, we introduce a complimentary compartment *S*^*w*^ for those who have received vaccines, but do not gain immunity. This class represents a non-existent but perceived immunity to the disease and as such we modify the model to account for a change in behaviour related to NPIs as a consequence. This compartment will also be a transient compartment for people who have waning immunity as there will be misalignment between when the protection from vaccine has diminished and when it has been perceived to have diminished.

To the best of our knowledge, no previous studies have studied the coupled effects of dynamic social distancing and cost-based relaxation, waning immunity, vaccination, and new variants of concern on the progression of the pandemic. Our study is organized as follows. In Section 2.1 we introduce the extended SEPIR model including new parameters, values for which are derived from existing literature or fit to data from Public Health Ontario (PHO) [36, 37]. We then present the estimated parameters using time horizons of public policy implementations in Ontario developed by Dick [38]. Additionally, we investigate the effect of waning immunity. We compare our results to publicly accessible data on positivity rate, daily incidence and seroprevalence [36, 39]. Following Moyles et al., [20], we focus our work on the Canadian province of Ontario. We discuss the conclusions of our work in Section 4.

## 2 Methods

### 2.1 SEPIR Model

We developed a compartmental mode based on the model proposed by Moyles et al. [20] which allows the various classes to change transmission dynamics through isolation and contact reduction. The Moyles et al. model is depicted in Figure 1, Panels (a) and (c). We extend the model to include vaccine-induced immunity, and perceived immunity (shown in Panel (b)) and waning immunity. Briefly, Panel (a) shows disease progression from susceptible (*S*) to recovered (*R*) through the different infection stages: non-infectious (*E*), pre-symptomatic infectious (*P*), asymptomatic infectious (*I*_*A*_), and symptomatic infectious (*I*_*S*_). Reported infections are denoted with subscript *M*. A natural waning immunity rate *ω*_*I*_ indicates the fraction of the recovered population that can once again become susceptible. Superscript *ξ* is shown in Panel (b) which illustrates the transition between individuals that are unvaccinated (*u*), vaccinated (*v*), or with perceived immunity (*w*). *p* is the rate of vaccination, *ω*_*V*_ is the waning rate from vaccine induced immunity, and *ω*_*W*_ it the waning rate of perceived immunity. Panel (c) illustrates movement between three social distancing classes, with subscripts 0, 1 and 2 denoting no social distancing, some social distancing and complete isolation, respectively. Individuals can move up and down the social distancing ladder. Note that the disease progression pathway shown in Panel A is the same for all individuals in different social distancing states (faded colours in Panel (a)). Movement between social distancing classes is allowed unless infection status is known (and requires full social distancing for all reported infections). Note that all infections transition from a susceptible state through to recovery but with rates and probabilities dependent on the immunity and social distancing status. As such, we denote our variables 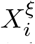 where *X* ∈ {*S, E, P, I*_*S*_, *I*_*A*_, *R*_*S*_, *R*_*A*_} is the disease state, the subscript *i* ∈ {0, 1, 2} is the physical distancing level, and the superscript *ξ* ∈ {*u, v, w*} indicates immunity status. The *M* subscript in panel (a) indicates those who have tested positive for the virus and are thus isolated from the population until recovery. We summarize each of the model disease classes as follows:

- Susceptible individuals denoted by 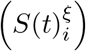, who are eligible to be infected by the pathogen.
- Exposed individuals denoted by 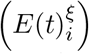, who have been infected but are incubating the virus. They are not transmissible and have a low enough viral load that they would not test positive for COVID-19.
- Pre-symptomatic individuals denoted by 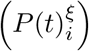, who are infectious but have not had the disease long enough to show symptoms.
- Infected-symptomatic individuals denoted by 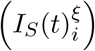 who are infectious and have started showing symptoms.
- Infected-asymptomatic individuals denoted by 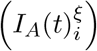, who are infectious and never show symptoms.
- Removed-symptomatic individuals denoted by 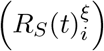, who were symptomatic, but are no longer infectious.
- Removed-symptomatic individuals denoted by 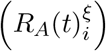, who were asymptomatic, but are no longer infectious,

**Figure 1:**
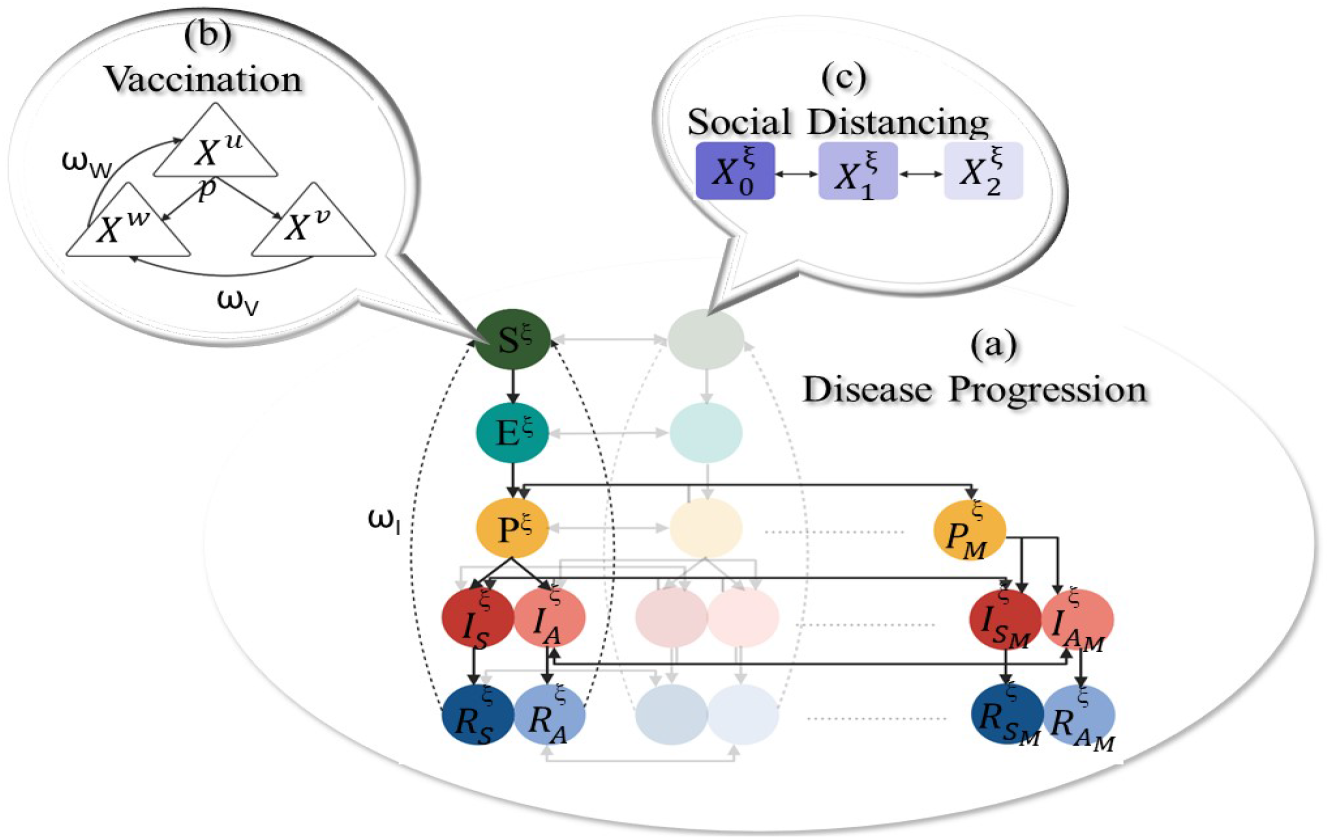
Schematic representation of the Susceptible- Exposed - Pre-symptomatic Infectious - Infectious Asymptomatic - Infectious Symptomatic - Recovered (SEPIR) with three levels of social distancing from no social distancing (subscript 0) to full isolation (subscript 2) (Panel (c)), and null, vaccine-induced or perceived immunity (Panel (b), superscript 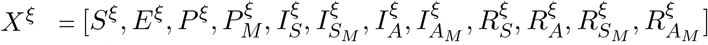. Known infections (via testing) are shown with subscript *M*.

with *t* as time in days since the onset of the pandemic, taken here to be March 10, 2020. For each of the population classes, we consider three levels of physical distancing: no isolation (subscript 0), partial isolation at contact reduction *δ* (subscript 1) and full isolation (subscript 2). All compartments sum to the total population, *N*, which is constant in time as we do not consider recruitment from birth or death. The governing differential equations for the full model depicted in Figure 1 are detailed in the Appendix.

### 2.2 COVID-19 Testing

In this study, we compare the number of cumulative and active reported infections, seroprevalence, daily incidence, and positivity rate calculated by our model with the data provided by Public Health Ontario.

#### Active reported infections

Active reported infections, *M*_*A*_ are defined by the sum of reported pre-symptomatic, asymptomatic, and symptomatic cases with different immunity levels who have not yet recovered, i.e. would not yield a negative test result. We define them as

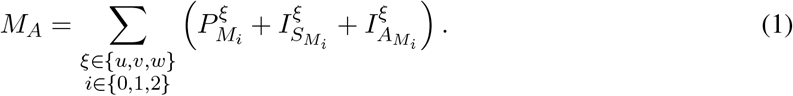

Note that we assume that all reported infections will fully isolate.

#### Cumulative reported infections

We define *M* to be the cumulative newly reported cases. We define the rate of change of cumulative reported incidence as a sum of pre-symptomatic, asymptomatic, and symptomatic infections who have tested positive at time *t* as follows

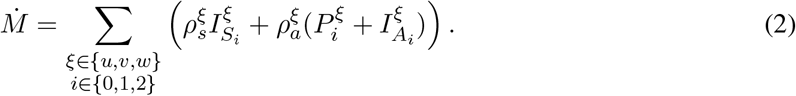

#### Total Vaccination Administered

Cumulative Vaccination, *V*_*A*_ is the total vaccines administered. The rate of change here is defined by the sum of all eligible vaccine recipients who vaccinate at time *t* with rate *p* and is defined as

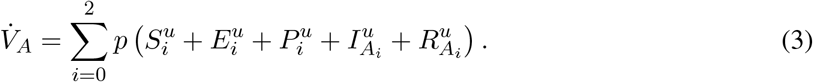

Importantly we assume that those who have symptomatic infection, have recovered from symptomatic infection, or have tested positive for having an infection are ineligible to receive a vaccine.

#### Seroprevalence

Serology testing, which tests someone’s blood to see if they have antibodies for COVID-19, is used as a measure of population-level infection and immunity. Seroprevalence, *S*_*R*_ is estimated by the number of people who test positive for COVID-19 antibodies based on serology data. Herein, we assume that people with COVID-19 antibodies will belong to the recovered class and thus

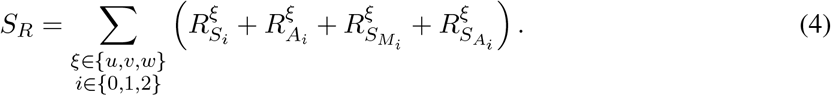

We note that individuals that wane out of the recovered classes will not have positive serology tests in our model.

#### Daily Reported Infection Incidence

*D*_*I*_ refers to the number of newly diagnosed COVID-19 cases per day, and is defined as

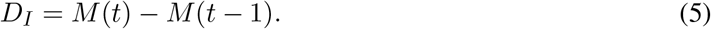

#### Positivity Rate

Since we assume that all individuals are eligible for testing then we can define the test positivity rate as the number of positive tests (daily incidence) divided by total tests administered across the entire population. We define the testing rate of symptomatic infections to be *ρ*_*s*_, and assume that the testing rate for all populations that are not infected or that have asymptomatic infection to have testing rate *ρ*_*a*_. Thus, we define the total tests *T*_*T*_

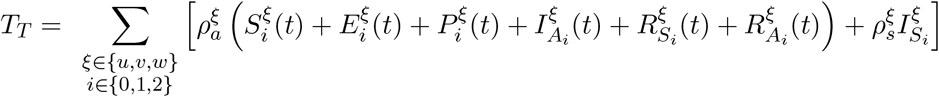

and the test positivity rate

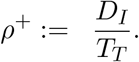

### 2.3 Physical distancing functions

We model the transition between the different social distancing classes as in [20], by assuming that individuals who are not vaccinated move from social distancing class 0 to class 1 with rate *μ*^*u*^ given by

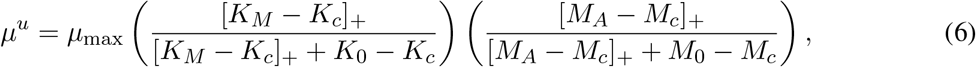

where *μ*_max_ is the maximal rate of social distancing, [·]_+_ = max(·, 0), and *K*_*M*_ is the doubling rate given by

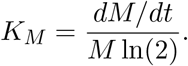

We assume that individuals transition from social distancing class 1 to class 2 with rate *μ*^*u*^*/*2 to take into account that people who have already reduced their contacts will be slower in fully isolating. Furthermore, we assume that individuals who are vaccinated are also slower in transitioning from social distancing class 0 to 1 by setting *μ*^*v*^ = *μ*^*w*^ = *μ*^*u*^*/*2. As we can see from the definition of the physical distancing function *μ*, the number of total and active reported cases determine if individuals will physical distance, and these two quantities are provided from testing.

Additionally, individuals decrease social distancing based on some cost, *C*, with rate *ν* defined as in [20],

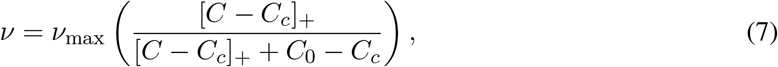

where *ν*_max_ is the maximal rate at which physical distancing can be relaxed. The cost of social distancing, primarily introduced by [20], is extended as follows

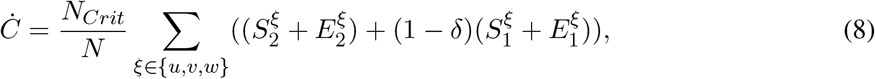

where the full cost occurs to those in all immunity groups who are susceptible or exposed (i.e. would test positive for the virus). As was done by Moyles et al. in [20], the cost is scaled to be in days where one day represents the cost of the entire population, *N*, fully or partially isolating. Individuals in the social distance class 1 have reduced transmissibility by a factor *δ*, and we assume this comes at a reciprocal burden cost of (1 − *δ*) per day.

### 2.4 Parameter Values and Estimation

In this study we estimate parameters (i) *K*_*c*_: critical approximate disease doubling rate to induce social distancing, (ii) *M*_*c*_: critical active cases to induce social distancing, and (iii) *ρ*_*a*_: testing rate for asymptomatic person to test positive as these parameters are assumed to vary within different public health mitigation periods. Additionally, we estimate *p*, the percentage of vaccinated people. We estimate these parameters in different time windows defined by the time period over which certain policies were in effect to investigate how their values change based on NPIs and pharmaceutical interventions. We choose the date and the category of the implemented NPI as developed by Dick et al. [38] where the authors used government resources and creditable news agencies to provide the timeline of categorized public health interventions from March 12, 2020, to January 5, 2022. In Table 2, we provide the dates of each time window and the corresponding policy.

For parameter fitting we use data from Public Health Ontario [36, 37] on cumulative and active reported cases, and total vaccines administered from March 10, 2020 to November 30, 2021. We start with an initial value for the first time window for the values of the parameters *K*_*c*_, *M*_*c*_, *ρ*_*a*_ and *p*, and then employ a non-linear least squares method to find the values of the parameters so that the simulated cumulative and active reported cases, and total number of vaccinations, best fit the data. For the second time window we use as initial value of the fitted values from the first time window, and estimate the values of the parameters again using the same fitting method. We repeat the same procedure until we have estimated the values of the parameters for all time windows.

The remaining model parameters are assigned the values listed in Tables 2 and in the Appendix.

### 2.5 Initial Conditions

We initialize all compartments to be zero except for the symptomatic infectious 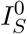 and the susceptible *S*^0^, assuming that 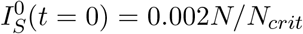 and *S*^0^(*t* = 0) = 0.98*N/N*_*crit*_, where *t* = 0 is the initial time and *N*_*crit*_ is the critical population at which healthcare resources are overwhelmed.

### 2.6 Sensitivity Analysis

We perform a sensitivity analysis on the waning parameters *ω*_*I*_, *ω*_*V*_, *ω*_*W*_ and the vaccine efficacy *ϵ*. To do so we generate 1000 samples of the parameters *ϵ, ω*_*I*_, *ω*_*V*_, and *ω*_*W*_ using the Latin hypercube method [40]. We assumed that the quickest vaccine induced immunity *ω*_*V*_ or infection induced immunity *ω*_*I*_, can wane is 4 months, and the slowest is 2 years [35]. The quickest the perceived induced immunity *ω*_*W*_ can wane is 4 months, and the slowest is 1 years [41]. We did not test the sensitivity of our model on the other parameters since our model is an extension of the model presented in [20] and the authors carried out sensitivity analysis on the model parameters. However, the waning parameters *ω*_*I*_, *ω*_*V*_, and *ω*_*W*_ and the vaccine efficacy *ϵ* are new parameters.

We take into account that the emergence of SARS-CoV-2 variants can affect transmission rate of the disease. If we define *β* as the transmission coefficient for the wild-type strain then when the Alpha variant (B.1.1.17) was dominant between February 15 and June 29, 2021 we modify the transmission coefficient to 1.5*β* accounting for the higher reproduction number of this variant [42]. Similarly, from June 29, 2021 to December 31, 2021 the Delta variant (B.1.617.2) was dominant and we modify the transmission to 2*β* [42]. For a subset of parameters, reasonable values were specified based on Health statistics, see Table 1 and 3 in the Appendix.

**Table 1:** The table shows what type of immunity each compartment has with 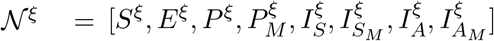 and 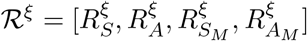, where *ξ* ∈ {*u, v, w*}.

| Classes | No immunity | Infection induced immunity | Vaccine induced immunity | Perceived induced immunity |
| --- | --- | --- | --- | --- |
| $\mathcal{N}^u$ | ✓ | | | |
| $\mathcal{R}^u$ | | ✓ | | |
| $\mathcal{N}^v$ | | | ✓ | ✓ |
| $\mathcal{R}^v$ | | ✓ | ✓ | ✓ |
| $\mathcal{N}^w$ | ✓ | | | ✓ |
| $\mathcal{R}^w$ | | ✓ | | ✓ |

**Table 2:** Estimated values of *K*_*c*_, *M*_*c*_, *ρ* and p.

| Time window | Important dates | | Rationale | $\beta$ | $K_c$ | $M_c$ | $\rho_a$ | $p$ |
| --- | --- | --- | --- | --- | --- | --- | --- | --- |
| 1 | 10-Mar-2020 | 07-Jun-2020 | Lockdown and gradual reopening | - | 0.0635 | 0.0239 | 0.0094 | - |
| 2 | 07-Jun-2020 | 20-Aug-2020 | Stage 2 and 3 mosaic | - | 0.0635 | 0.0239 | 0.0061 | - |
| 3 | 20-Aug-2020 | 25-Dec-2020 | Tightening Measures, second wave | - | 0.0160 | 0.0000 | 0.0021 | - |
| 4 | 25-Dec-2020 | 19-Jan-2021 | Tightening Measures, second wave | - | 0.0000 | 0.2885 | 0.0024 | - |
| 5 | 19-Jan-2021 | 28-Jan-2021 | Stay at home | - | 0.0000 | 0.0000 | 0.0024 | - |
| 6 | 28-Jan-2021 | 15-Feb-2021 | Stay at home | - | 0.0031 | 0.0000 | 0.0038 | - |
| 7 | 15-Feb-2021 | 12-Mar-2021 | Reopening scenarios | $1.5\beta$ | 0.0048 | 0.0000 | 0.0040 | 0.0011 |
| 8 | 12-Mar-2021 | 04-Apr-2021 | Reopening scenarios | $1.5\beta$ | 0.0070 | 0.0000 | 0.0020 | 0.0000 |
| 9 | 04-Apr-2021 | 09-May-2021 | Emergency stay at home | $1.5\beta$ | 0.0000 | 0.3312 | 0.0024 | 0.0002 |
| 10 | 09-May-2021 | 10-Jun-2021 | Emergency stay at home | $1.5\beta$ | 0.0000 | 0.0000 | 0.0029 | 0.0016 |
| 11 | 10-Jun-2021 | 15-Jun-2021 | S1 | $1.5\beta$ | 0.0096 | 0.0097 | 0.0040 | 0.0187 |
| 12 | 15-Jun-2021 | 29-Jun-2021 | S1 | $1.5\beta$ | 0.0096 | 0.0097 | 0.0043 | 0.0153 |
| 13 | 29-Jun-2021 | 15-Jul-2021 | S2 | $2\beta$ | 0.0096 | 0.0097 | 0.0010 | 0.0288 |
| 14 | 15-Jul-2021 | 01-Sep-2021 | S3 | $2\beta$ | 0.0096 | 0.0097 | 0.0003 | 0.0221 |
| 15 | 01-Sep-2021 | 30-Nov-2021 | S3 | $2\beta$ | 0.0096 | 0.0097 | 0.0020 | 0.0230 |

## 3 Results

### 3.1 Time windows

We provide the fitting results for each time window in Table 2. From the start of the pandemic until August 20, 2020 (time windows 1 and 2), the values of the parameters *K*_*c*_ and *M*_*c*_ remain the same indicating that individuals had the same level of vigilance during that time period, while the testing rate *ρ*_*a*_ is high during the first time window, but it decreases during the second time window. The decrease in the values of *K*_*c*_ and *M*_*c*_ between August 20 and December 25, 2020 shows that individuals became more cautious, while testing decreases further compared to the previous time period. During that time window, more strict measures were implemented in Ontario explaining the increased vigilance. During time window 4, we observe that the value of *K*_*c*_ drops, but the values of *M*_*c*_ and *ρ*_*a*_ increase. Increase in the value of *M*_*c*_ indicates that more cases are needed to induce social distancing, but the critical doubling rate is zero meaning that any increase in the doubling rate leads to more vigilance. Increase in the value of *M*_*c*_ and reduction in the value of *K*_*c*_ might occur during the exponential phase of spread of the disease when the number of cases might not be high, but the doubling rate is high and individuals are more cautious knowing that the number of cases is exponentially growing. From January 19 to February 15, 2021 (time windows 5 and 6), a stay-at-home order was in effect in Ontario, and this resulted in people increasing social distancing as the value of *M*_*c*_ remains zero implying that any number of cases triggers social distancing. From February 15 to April 4, 2021 (time windows 7 and 8), although the government was considering relaxation of the mitigation measures, the values of *M*_*c*_ remain zero showing that individuals were still vigilant and continue to social distance if the number of cases is non-zero. During time windows 9 and 10, the stay-at-home order was again in effect. Although, the value of *M*_*c*_ increased during time window 9, the value of *K*_*c*_ remains zero for both time windows indicating that people are reducing their contacts if the doubling rate is greater than zero. Finally, from June 10 to November 30, 2021 (time windows 11 to 15), the values of *K*_*c*_ and *M*_*c*_ remain constant and increase compared to the time period between May 9 and June 15. It is possible that the increase in vaccine coverage resulted in people being more relaxed about social distancing, and would reduce their social activities only when the doubling rate or number of cases would surpass the value of *K*_*c*_ and *M*_*c*_, respectively.

### 3.2 Waning immunity

In Figure 2 we present the results from our sensitivity analysis on the daily minimum and maximum active cases given by the 1000 samples of the parameters *ϵ, ω*_*I*_, *ω*_*V*_, *ω*_*W*_ and the estimated values of *K*_*c*_, *M*_*c*_, *ρ* and *p*. The values for *ω*_*I*_ and *ω*_*V*_ are chosen on a per-day basis, and We observe that the minimum and maximum number of daily active cases are similar in magnitude up to December 25, 2020, which implies that model predictions are not affected by the waning immunity parameters up to this date. The minimum number of cases (blue line) corresponds to values of the parameters *ω*_*I*_ between 0.0013 and 0.0016, and *ω*_*V*_ between 0.0015 and 0.0020, (both very close to the assumed two-year upper bound) we can see that the model predicts no infection after July 29, 2021.. This shows that if we assume slow waning rates, the model does not capture the fourth wave, meaning that the number of individuals left in the susceptible compartment is not sufficiently large for the disease to spread. On the other hand, the model overestimates the number of active cases after July 29, 2021, if we assume that the infection and vaccine induced immunity fade quickly (red line) with *ω*_*V*_ ∈ (0.0075, 0.0082) and *ω*_*I*_ ∈ (0.0062, 0.0074) near the assumed 4-month lower bound in waning time.

**Figure 2:**
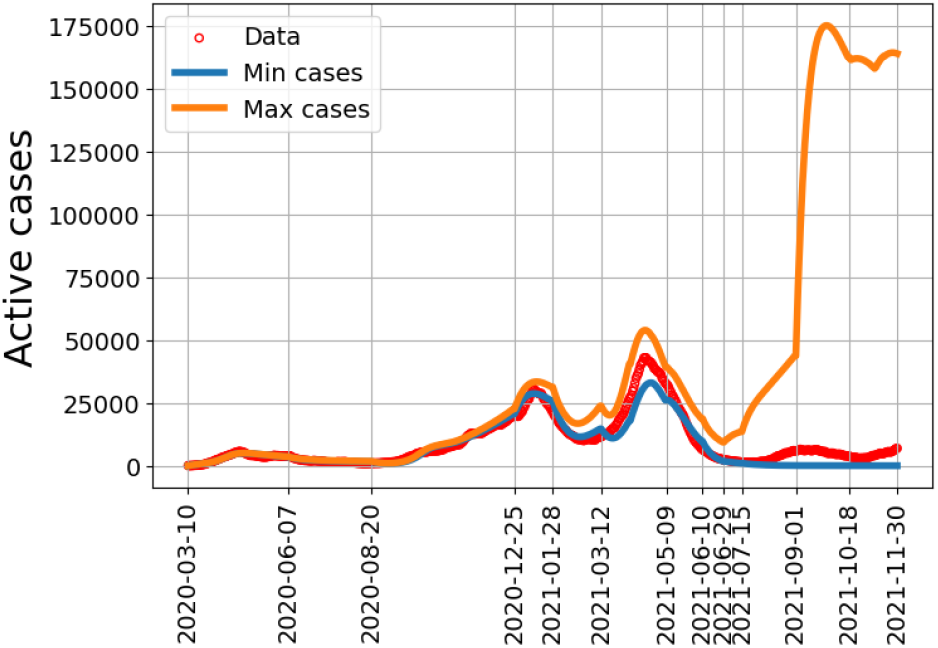
The minimum and maximum number of active cases per day, obtained by numerically solving the model equations, for different values of *ϵ, ω*_*I*_, *ω*_*V*_ and *ω*_*W*_.

### 3.3 Model Prediction Vs. Observed Data

The results from the model fitting of *K*_*c*_, *M*_*c*_, *ρ*_*a*_ and *p* are illustrated in Figure 3. Here, we also plot data on cumulative reported infections (top panel), active reported infections (middle panel), and the total vaccines administered (bottom panel) [37]. We observe a satisfactory model prediction of observed data for cumulative incidence and total vaccination administered criteria between March 10, 2022, and November 30, 2021. For active reported infections, the fit is satisfactory until August 2021 after which there is an overshoot compared to the data.

**Figure 3:**
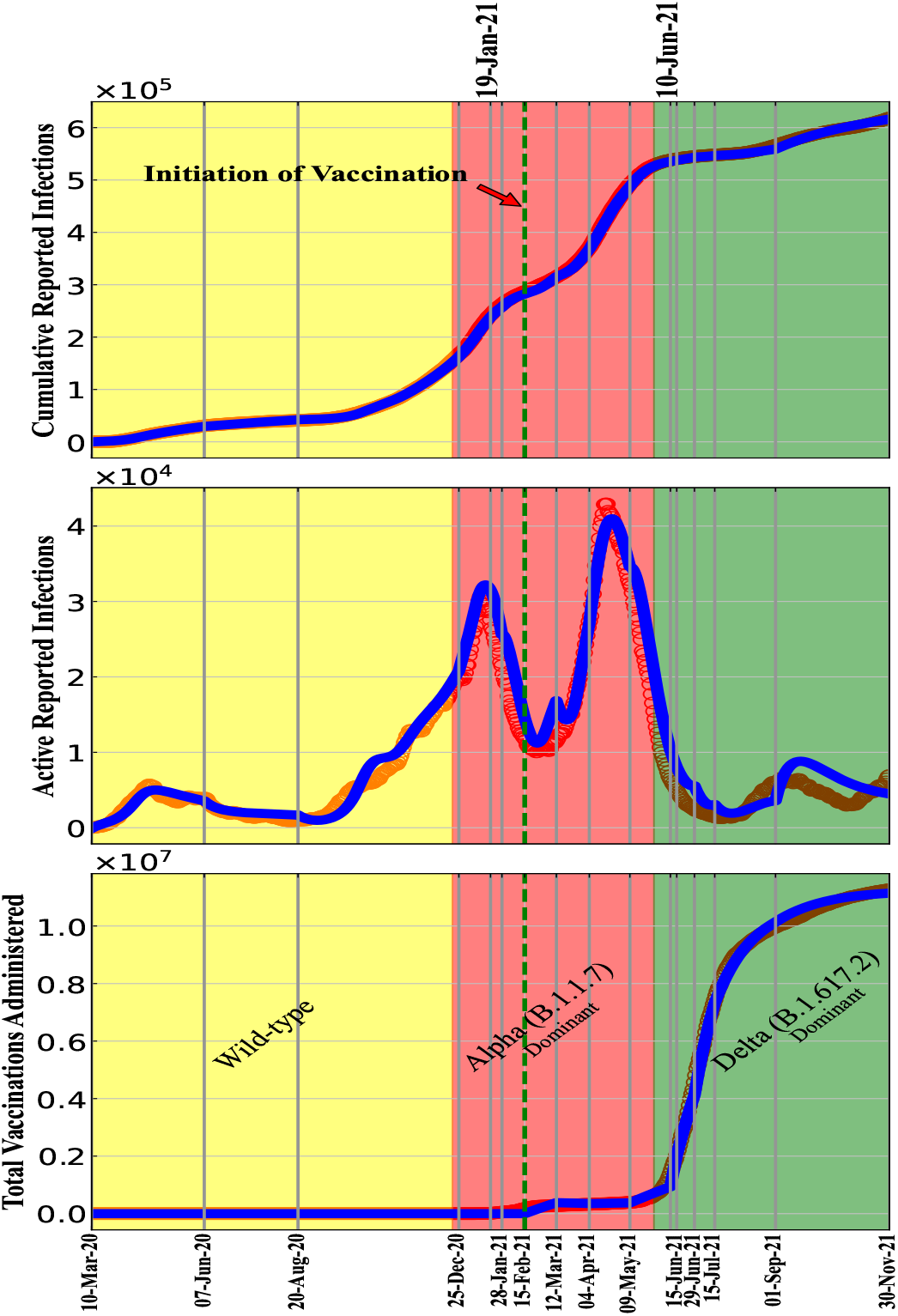
Comparison between model simulations and Ontario Data Catalogue. Model predictions fits to data from [36] in panel (a) and (b), and fits to data from [37] in panel (c), from March 10 2020 to November 30 2021. The green vertical dashed line shows the vaccination starting date. In the red (green) shade area the diseases transmission rate of the variants, was assumed to be one and half times (double) greater than the transmission rate of wild type.

Further, we compare the model results with data which was not used to fit the model, particularly seroprevalence, daily incidence, and positivity rate. The evolution of model predicted seroprevalence in different cohorts, daily incidence, positivity rate, and the corresponding health data are depicted over time in Figure 4. Although, there is a relatively good agreement between data and simulation for daily incidence and positivity rate, the estimated seroprevalence is higher than the data suggest. The data on seroprevalence are based on studies from blood donors aged 16+ from Canadian Blood Services (CBS) [39, 43]. We note here that our model does not distinguish between serology and T-cell mediated immunity whereas the CBS data report the results from serological testing only. It is possible for individuals to have T-cell immunity even when antibody levels have waned. A recent study reported that high T-cell memory levels can protect against COVID-19 infection [44]. Additionally, we assume a well-mixed population in our model. This can also increase our estimates as we do not include contact networks. Finally, our model presents immunity from the entire population irrespective of age whereas the CBS serological testing is conducted in ages 16+ only. The inclusion of age structure and contact matrices may reduce our seroprevalence estimates. However, even with age-structuring Dick et al. estimated seroprevalence higher than suggested by the data, although less of a difference than we see here [35] Given these points we find that it is not surprising that our model and the data do not agree. In future, we will consider a model with age-structure to see if this will provide immunity estimates closer to the serological testing data.

**Figure 4:**
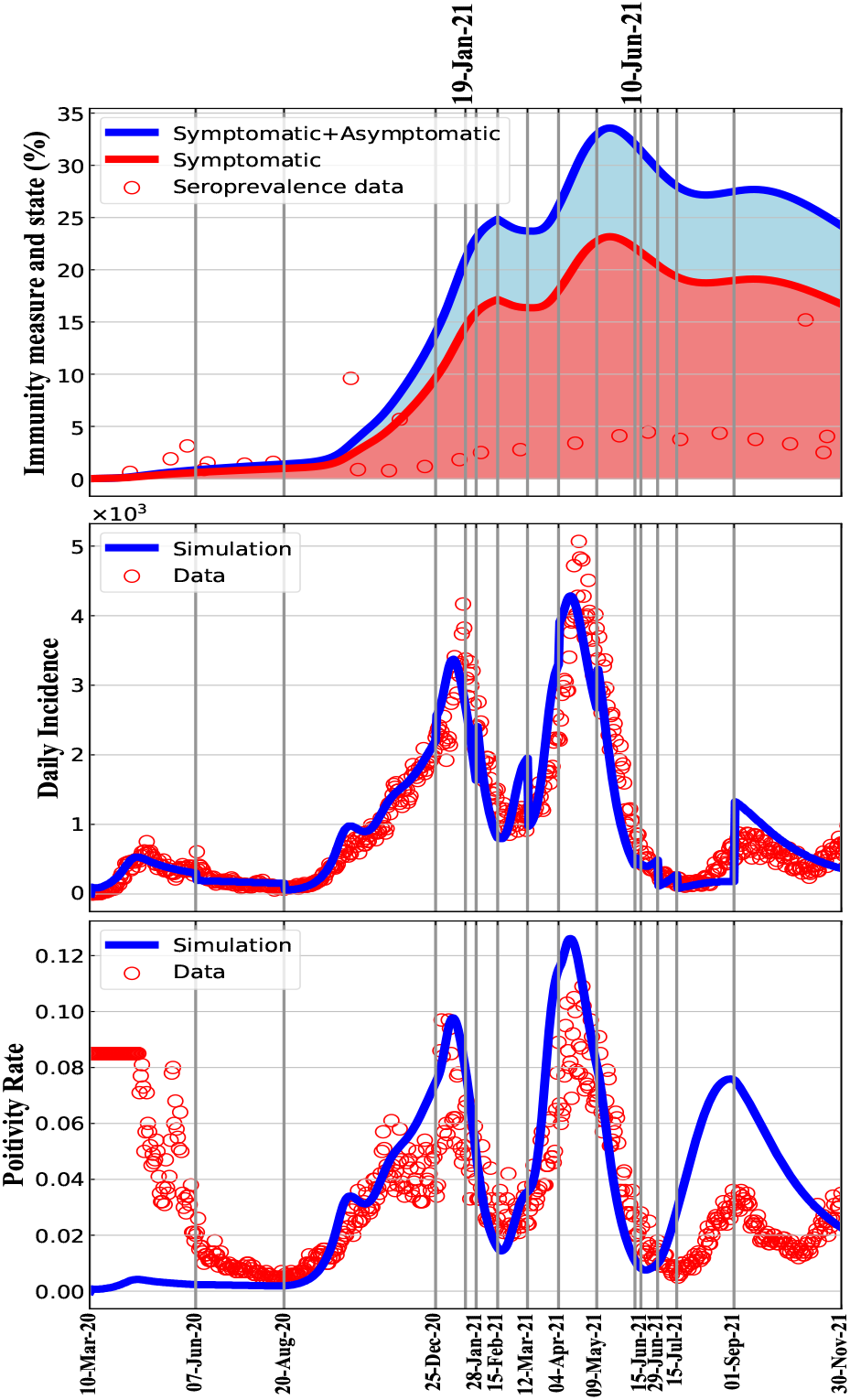
The simulation results for the seroprevalence, daily incidence and positivity rate have been projected with health data from March 10 2020 to November 30, 2021.

## 4 Discussion

In this work, we proposed a compartmental model coupling the effects of dynamic social distancing and cost-based relaxation, different immunity levels, vaccination, and new variants of concern to study which can recreate the history of the COVID-19 pandemic up to December 2021 in Ontario. The model can predict different quantities of interest including active cases, vaccination, daily incidence and positivity rate. However, our model predictions on the seroprevalence are different from the data which could be due to the challenges on estimating population seroprevalence from serological testing and/or the homogeneous mixing assumption for our model.

We concluded that if we assume that it takes 2 years for disease or vaccine induced immunity to wane, our model does not capture the fourth wave in Ontario. Our sensitivity analysis showed that waning immunity would not change anything in the model predictions on active cases until December 25, 2020. However, the model is more sensitive after December 25, 2020, and the values of the waning parameters affect the fitting results. Our simulations and sensitivity analysis showed that waning immunity is crucial to capture more accurately the disease dynamics and predict multiple waves over a long time period. In future work, we want to extend the time period to include the Omicron variant and study the effect of evading immunity on disease dynamics.

We estimated key parameters affecting the vigilance of individuals at different time windows and found that NPIs influence how they increase or decrease their contacts. For example, they are more cautious when stricter measures are introduced such as stay-at-home orders or lockdowns. Our results also indicate that people started being more relaxed about social distancing after May 2021, which is approximately when vaccine coverage increased in Canada. This shows the importance of having a model that incorporates dynamic human behaviour in order to capture how people change their behaviour based on the disease dynamics and NPIs.

As a case study, we used different health data from Ontario to evaluate our model predictions. However, our modeling framework can be easily adapted to any other country or province for which relevant data are available. Our modelling approach can provide important insights how NPIs and vaccination can influence the health decisions people make during epidemics, and better understand how disease dynamics are affected by those decisions.

## Data Availability

All data produced are available online at: https://data.ontario.ca/en/dataset/status-of-covid-19-cases-in-ontario, https://data.ontario.ca/en/dataset/covid-19-vaccine-data-in-ontario, and https://www.covid19immunitytaskforce.ca/seroprevalence-in-canada/.

## CRediT authorship contribution statement

**Jeta Molla:** Conceptualization, Methodology, Software, Visualization, Writing – original draft, Writing – review & editing. **Suzan Farhang-Sardroodi:** Conceptualization, Methodology, Software, Visualization, Data Curation, Investigation. **Iain R Moyles:** Conceptualization, Methodology, Software, Funding acquisition, Supervision, Writing – original draft, Writing – review & editing. **Jane M Heffernan:** Conceptualization, Methodology, Funding acquisition, Supervision, Writing – review & editing.

## Acknowledgments

This work was supported by NSERC (JMH, IM), CIHR (JMH), and the OMNI-REUNI NSERC-PHAC Emerging Infectious Disease Modelling Initiative (JMH, IM). JM would like to acknowledge support from an OMNI-REUNI postdoctoral fellowship.

## Appendix

### Sensitivity analysis

In Figure 5 we present the scatterplots for the estimated parameters *K*_*c*_, *M*_*c*_, *ρ* and *p* versus the waning rates *ω*_*I*_, *ω*_*V*_, and *ω*_*W*_. The results show that there is no relationship between *K*_*c*_, *M*_*c*_, *ρ* and *ω*_*I*_, *ω*_*V*_, *ω*_*W*_. The waning rates *ω*_*I*_, *ω*_*V*_ and *ω*_*W*_ determine how fast the infection or vaccine-induced immunity, and the perceived-induced immunity wane, and it would be expected that they do not influence the values of the behaviour parameters *ρ, M*_*c*_, *K*_*c*_ since individuals do not know when their immunity wanes. While there is no relationship between *p* and *ω*_*I*_, the results indicate a negative relationship of moderate strength between *p* and *ω*_*V*_, and *p* and *ω*_*W*_. This implies that as the rate at which individuals transition from *u* to *w* decreases, the vaccination rate has to increase to fit the vaccination data.

**Figure 5:**
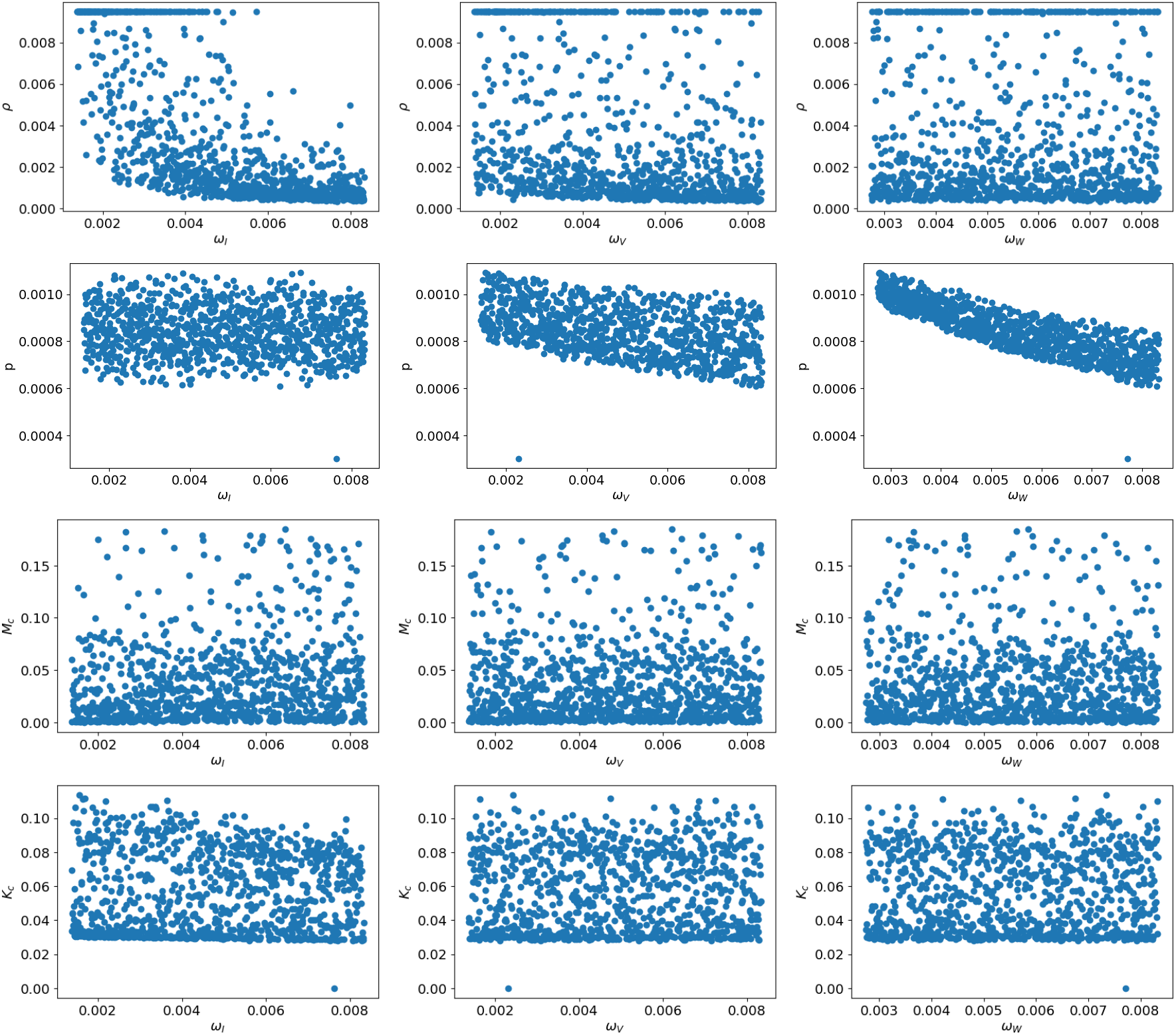
Scatterplots of the parameters *K*_*c*_, *M*_*c*_, *ρ, p* versus *ω*_*I*_, *ω*_*V*_ and *ω*_*W*_.

### Differential Equation Models

The differential equation models for people with natural immunity (Eq: A.9), vaccine/perceived induced immunity (Eq: A.10) and perceived induced immunity (Eq: A.11) are given by the following equations.

#### 4.1 Natural Immunity

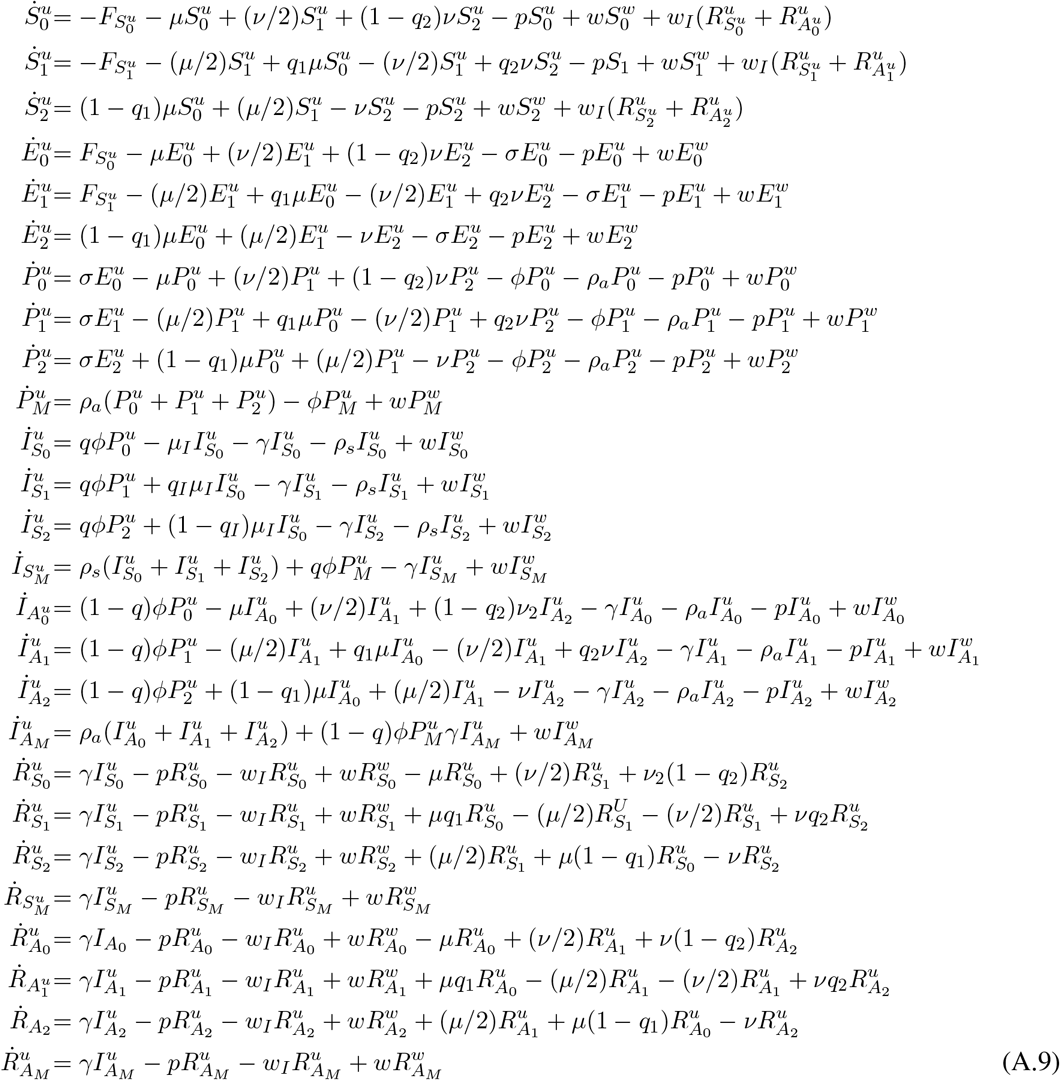

#### 4.2 Vaccine/Perceived Induced Immunity

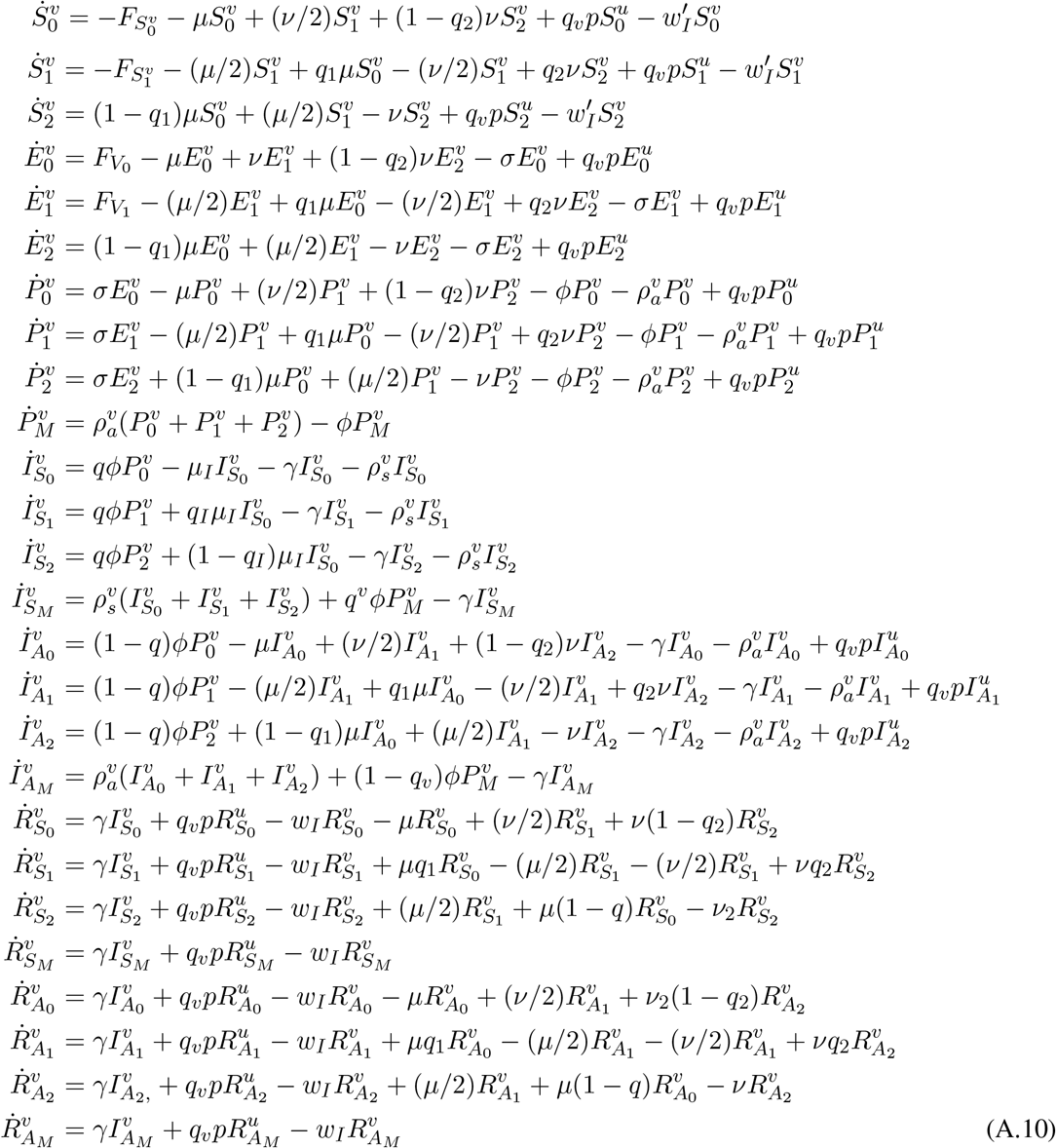

### Perceived Induced Immunity

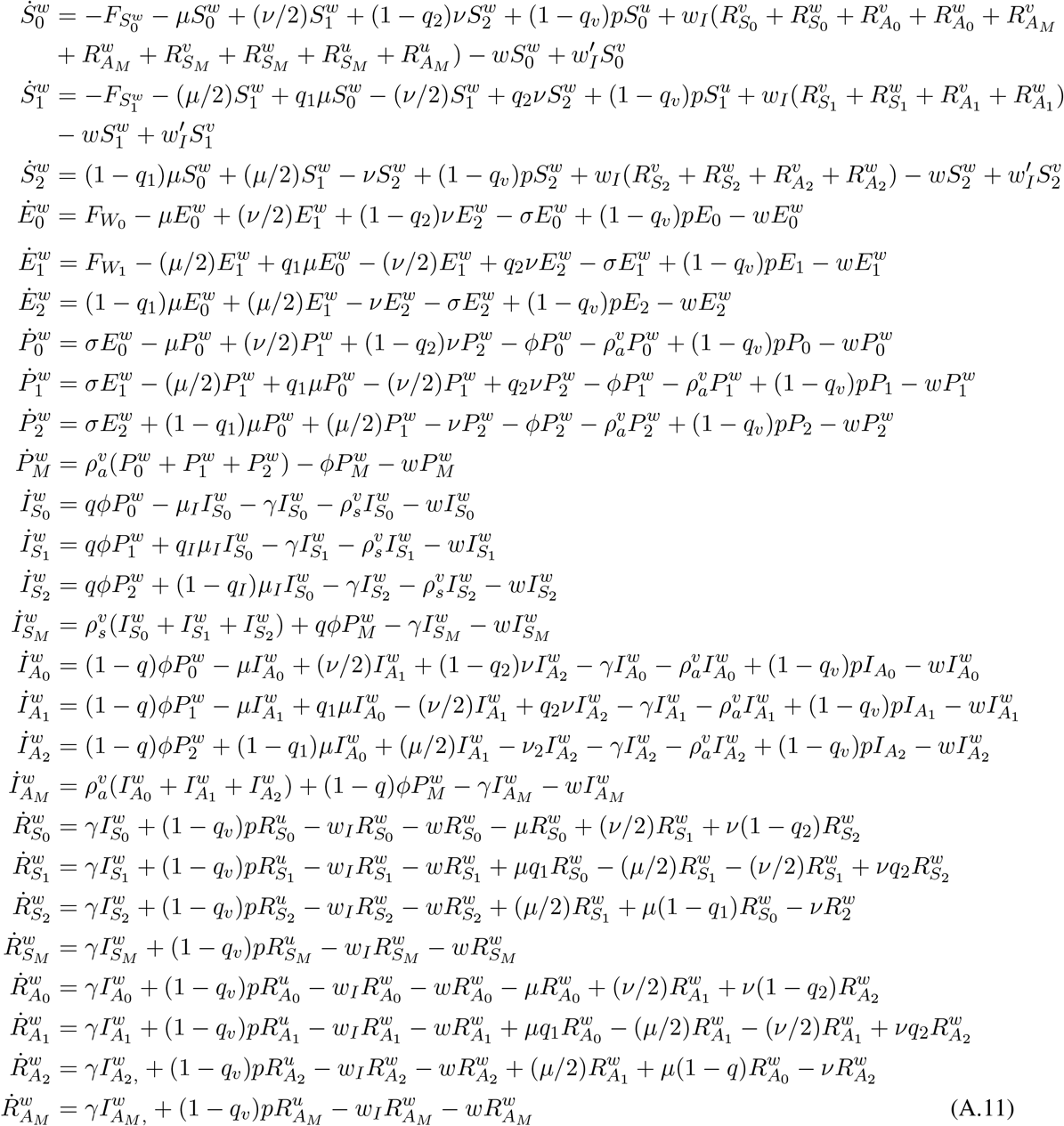

Where 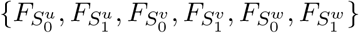 are force of infections and defined in Table 3.

**Table 3:** Force of Infections

| Infectious |  |
| --- | --- |
| $(I_{S_0}^u + I_{S_0}^v + I_{S_0}^w) + \alpha(P_0^u + I_{A_0}^u + P_0^v + I_{A_0}^v + P_0^w + I_{A_0}^w + \delta(P_1^u + I_{A_1}^u + P_1^v + I_{A_1}^v + P_1^w + I_{A_1}^w)) + \delta(I_{S_1}^u + I_{S_1}^v + I_{S_1}^w)$ | |
| $F_{\xi_0} = \frac{N_{crit}\beta\xi_0}{N}(\text{Infectious}), \xi_0 = S_0^u, S_0^w, \varepsilon S_0^v$ | $F_{\xi_1} = \frac{N_{crit}\delta\beta\xi_1}{N}(\text{Infectious}), \xi_1 = S_1^u, S_1^w, \varepsilon S_1^v$ |

**Table 4:** Values of the model parameters.

| Parameter | Definition | Value | Reference |
| --- | --- | --- | --- |
| $R_0$ | Basic reproduction number | 2.4 | [20] |
| $\beta$ | Transmission rate | 0.223 | Calculated |
| $\sigma$ | Latent period | 2 days<sup>-1</sup> | [20] |
| $\phi$ | Pre-symptomatic period | 4.6 days<sup>-1</sup> | [20] |
| $\gamma$ | Infectious period | 10 days<sup>-1</sup> | [20] |
| $\delta$ | Reduction in transmission due to social distancing in class 1 | 0.25 | Chosen |
| $\alpha$ | Reduction in transmission due to being asymptomatic | 0.5 | Chosen |
| $Q$ | Proportion of infected individuals who show symptoms | 0.69 | Median value |
| $\mu_{\max}$ | Maximal rate at which an un-vaccinated individual transitions from a less socially distant class to a more socially distant class | 1 days<sup>-1</sup> | Chosen |
| $\mu_{\max}^v, \mu_{\max}^w$ | Maximal rate at which a vaccinated individual transitions from a less socially distant class to a more socially distant class | 0.5 days<sup>-1</sup> | Chosen |
| $\nu_{\max}$ | Maximal rate at which an un-vaccinated individual moves from a more socially distant class to a less socially distant class | 1 days<sup>-1</sup> | Chosen |
| $\nu_{\max}^v, \nu_{\max}^w$ | Maximal rate at which a vaccinated moves from a more socially distant class to a less socially distant class | 2 days<sup>-1</sup> | Chosen |
| $\mu_I$ | Rate at which people showing symptoms choose to isolate | 0.01 days<sup>-1</sup> | Chosen |
| $q_0$ | Proportion of $S_0$ socially distancing into $S_1$ | 0.9 | Chosen |
| $q_2$ | Proportion of $S_2$ relaxing social distancing into $S_1$ | 0.6 | Chosen |
| $q_I$ | Proportion of symptomatic individuals $I_{S_0}$ who isolate into $I_{S_1}$ | 0.6 | Chosen |
| $C_c$ | Critical cost to induce social relaxation | 50 days | Chosen |
| $C_0$ | Cost that leads to half the maximal rate of social relaxation | 100 days | Chosen |

**Table 5:** Model Parameters

| Fixed parameters |  |  |  |
| --- | --- | --- | --- |
| parameter | Definition | Value | comment |
| $q_v$ | $(1 - q_v)$ is the fraction of people with perceived induced immunity | 1 | Chosen |
| $\rho_a^v$ | Testing rate of vaccinated people | $0.5\rho_a$ | Chosen |
| $\varepsilon$ | $(1 - \varepsilon)$ is the efficacy of the vaccine | 0.0 | Chosen |
| $\omega_W$ | Media waning rate | 0.07 | Chosen |
| $\omega_I$ | Disease waning rate | 0.005 | Chosen |
| $\omega_V$ | Vaccine waning rate | 0.005 | Chosen |

